# Left ventricular function and clinical outcomes according to vericiguat and sacubitril/valsartan use in heart failure with nonpreserved ejection fraction: a real-world study

**DOI:** 10.64898/2026.01.29.26345176

**Authors:** Haruyuki Kinoshita

**Affiliations:** Department of Cardiology, NHO Kure Medical Center, Kure, Japan

**Author notes:** Corresponding Author: Haruyuki Kinoshita, MD, Department of Cardiology, NHO Kure Medical Center, Kure, Japan, Aoyamacho 3-1, Kure 737-0023, Japan. Funding: This research received no external funding. Ethical Approval: Approved by NHO Kure Medical Center Ethics Review Committee No. 2023-43.

**Keywords:** heart failure, vericiguat, sacubitril/valsartan, heart failure with nonpreserved ejection fraction, add vericiguat, composite endpoint

## Abstract

**Background:** Vericiguat and sacubitril/valsartan both modulate the nitric oxide–soluble guanylate cyclase–cyclic guanosine monophosphate signaling pathway and may improve myocardial function in patients with heart failure.

**Objectives:** To compare the effects of vericiguat, sacubitril/valsartan, and their combination on left ventricular function and clinical outcomes in heart failure with nonpreserved ejection fraction patients.

**Methods:** In this retrospective real-world study, patients were classified into three groups: vericiguat added to guideline-directed medical therapy (vericiguat group), sacubitril/valsartan-based therapy (sacubitril/valsartan group), and combined sacubitril/valsartan plus vericiguat therapy (add-vericiguat group). Changes in left ventricular ejection fraction (ΔLVEF), in stroke volume (ΔSV), and in log-transformed N-terminal pro–B-type natriuretic peptide (ΔLog_10_ NT-pro BNP) from baseline to 1 year were evaluated. Clinical outcomes were also assessed.

**Results:** At 1 year, LVEF significantly improved in both the vericiguat group (p = 0.02) and the sacubitril/valsartan group (p < 0.001). There was no significant difference in ΔLVEF between these two groups (p = 0.25). In contrast, the add-vericiguat group demonstrated a significantly greater improvement in ΔLVEF compared with the vericiguat group alone (p = 0.01).

**Conclusions:** In a real-world setting, vericiguat was associated with improvements in left ventricular function comparable to those of sacubitril/valsartan, and combination therapy provided incremental benefits. Vericiguat may serve as an alternative or adjunctive treatment option, particularly in patients unable to tolerate or maintain angiotensin receptor–neprilysin inhibitor therapy.

## Introduction

A participant-level pooled analysis of PARADIGM-HF^1^ and PARAGON-HF^2^ reported a relative 16% risk reduction in the efficacy of sacubitril/valsartan compared with enalapril for the composite endpoint of cardiovascular mortality and hospitalization for heart failure in patients with a left ventricular ejection fraction (LVEF) of 32.5–52.5%.^3^

On the other hand, the efficacy of vericiguat has been recognized based on evidence from the VICTORIA trial.^4^

In Japan, vericiguat has been introduced relatively flexibly to patients with heart failure with reduced ejection fraction (HFrEF) who are already receiving standard cardioprotective agents, such as sacubitril/valsartan, beta-blockers, and mineralocorticoid receptor antagonists (MRAs), and who are considered at risk of worsening heart failure.^5^ Furthermore, given its pharmacological mechanism, vericiguat use has been increasing in Japan with the expectation of improving reverse remodeling and long-term outcomes.^6^ At our institution, vericiguat has been used in patients receiving sacubitril/valsartan since 2021. Preliminary real-world data from a subset of this cohort suggested a potential association between the addition of vericiguat and clinical composite endpoints.^7^

However, whether vericiguat confers additional benefits on cardiac remodeling beyond sacubitril/valsartan remains unclear. Therefore, the present study aimed to evaluate the impact of vericiguat on cardiac remodeling according to heart failure stage in patients with heart failure and nonpreserved ejection fraction treated with sacubitril/valsartan, focusing on longitudinal changes in left ventricular ejection fraction, stroke volume, and log_10_ NT-proBNP.

Secondary analyses explored associations with clinical outcomes using time-to-event analyses.

## Methods

This study utilized the same institutional registry as in our previous report published in the European Journal of Heart Failure (EJHF) (Kinoshita et al., 2025).^7^ However, the current analysis focused on different aspects of the cohort, particularly the echocardiographic parameters corresponding to Figures 1–5 and Table 1 in the previous publication, which have not been previously analyzed or discussed in detail.

**Figure 1:**
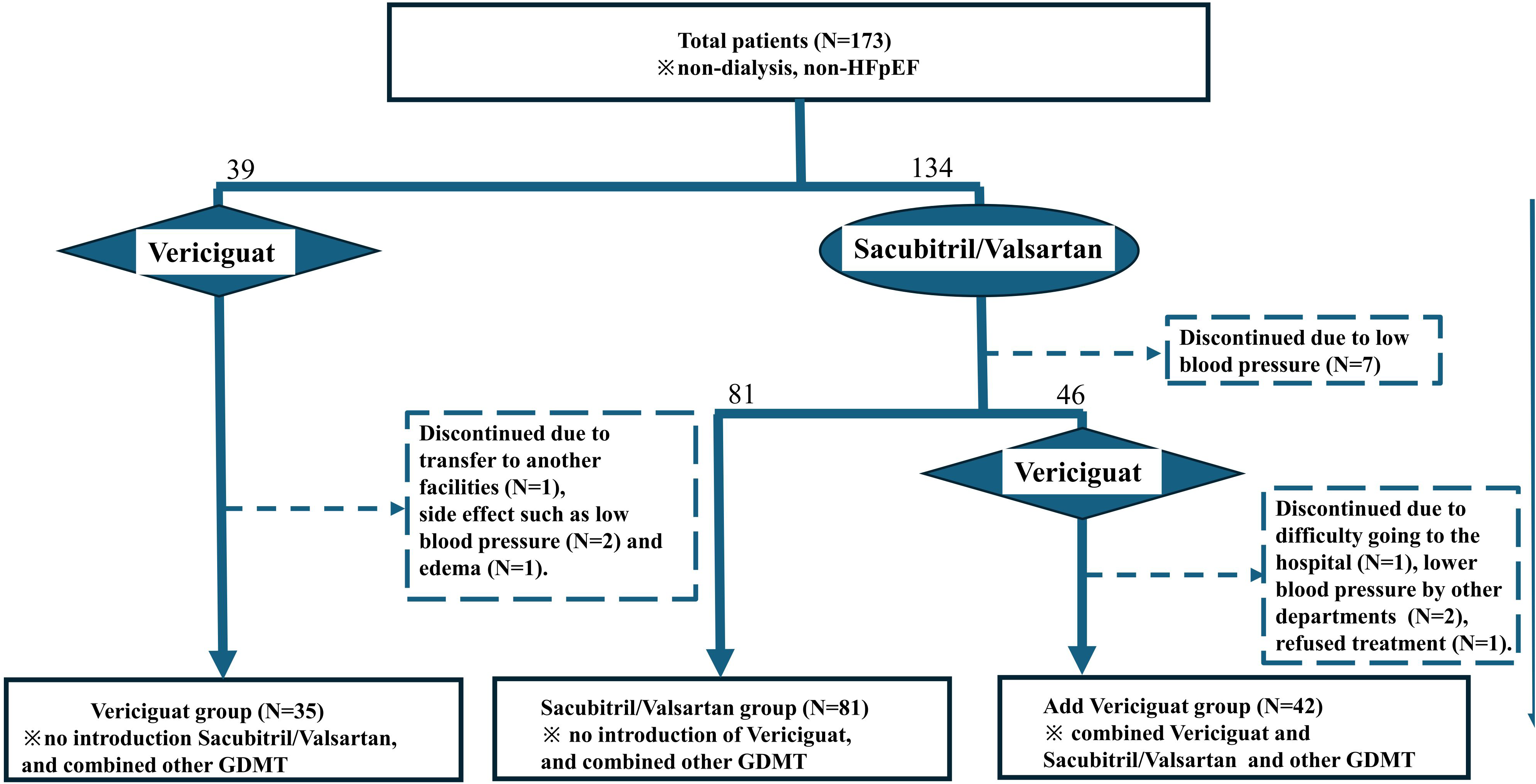
Illustration of the study protocol. In total, 173 non-dialysis patients without HFpEF underwent pharmacological treatment for heart failure. Thirty-nine patients received vericiguat in combination with standard heart failure medications other than sacubitril/valsartan, including ACE-I or ARBs, β-blockers, MRAs, SGLT-2 inhibitors, and diuretics. One patient discontinued vericiguat because of transfer to another hospital, two discontinued vericiguat because of hypotension, and one discontinued vericiguat because of lower leg edema. Thirty-five patients in the vericiguat group were followed up. Sacubitril/valsartan was initiated in 134 patients, along with β-blockers, MRA, SGLT-2 inhibitors, or diuretics. Seven of these patients discontinued treatment owing to hypotension and were excluded from the study. Eighty-one patients continued treatment without vericiguat; this group was referred to as the sacubitril/valsartan group. In addition to sacubitril/valsartan. One patient was excluded because he was unable to visit the hospital. Two were managed by other departments, but vericiguat was discontinued because of hypotension. One patient was excluded because of a refusal to take oral medication due to dementia. Ultimately, 42 patients were treated and followed up in the vericiguat group. Ultimately, 158 patients were included in this study.

**Figure 2:**
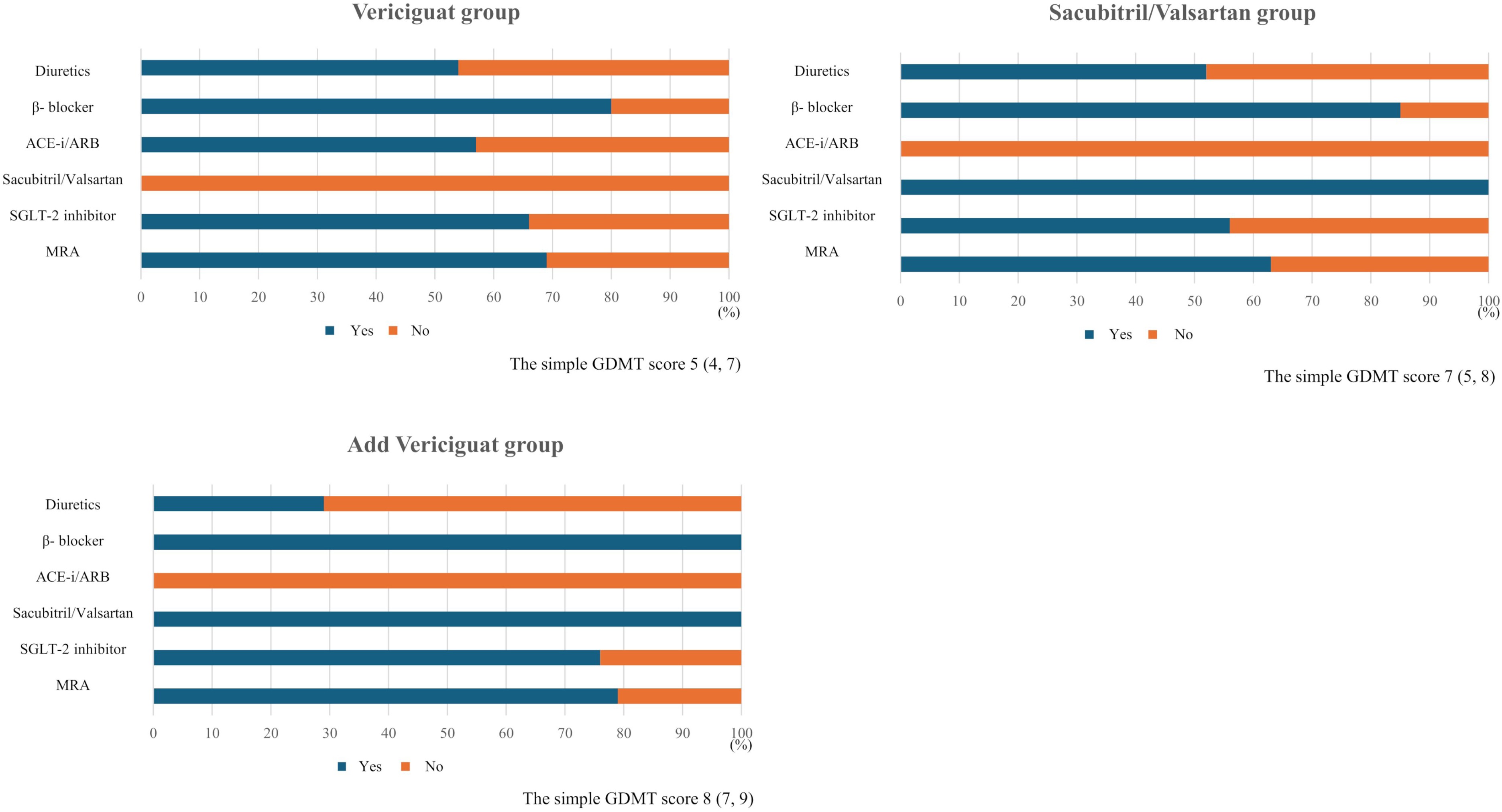
Prescription rates of standard heart failure medications in 158 patients. Medication use and median GDMT scores in each treatment group. The proportions of patients receiving diuretics, β-blockers, ACE inhibitors or ARBs, sacubitril/valsartan, SGLT2 inhibitors, and MRAs were 54%, 80%, 57%, 0%, 66%, and 69% (vericiguat group); 52%, 85%, 0%, 100%, 56%, and 63% (sacubitril/valsartan group); and 29%, 100%, 0%, 100%, 76%, and 79% (add-vericiguat group), with median simple GDMT scores of 5, 7, and 8 points.

**Figure 3:**
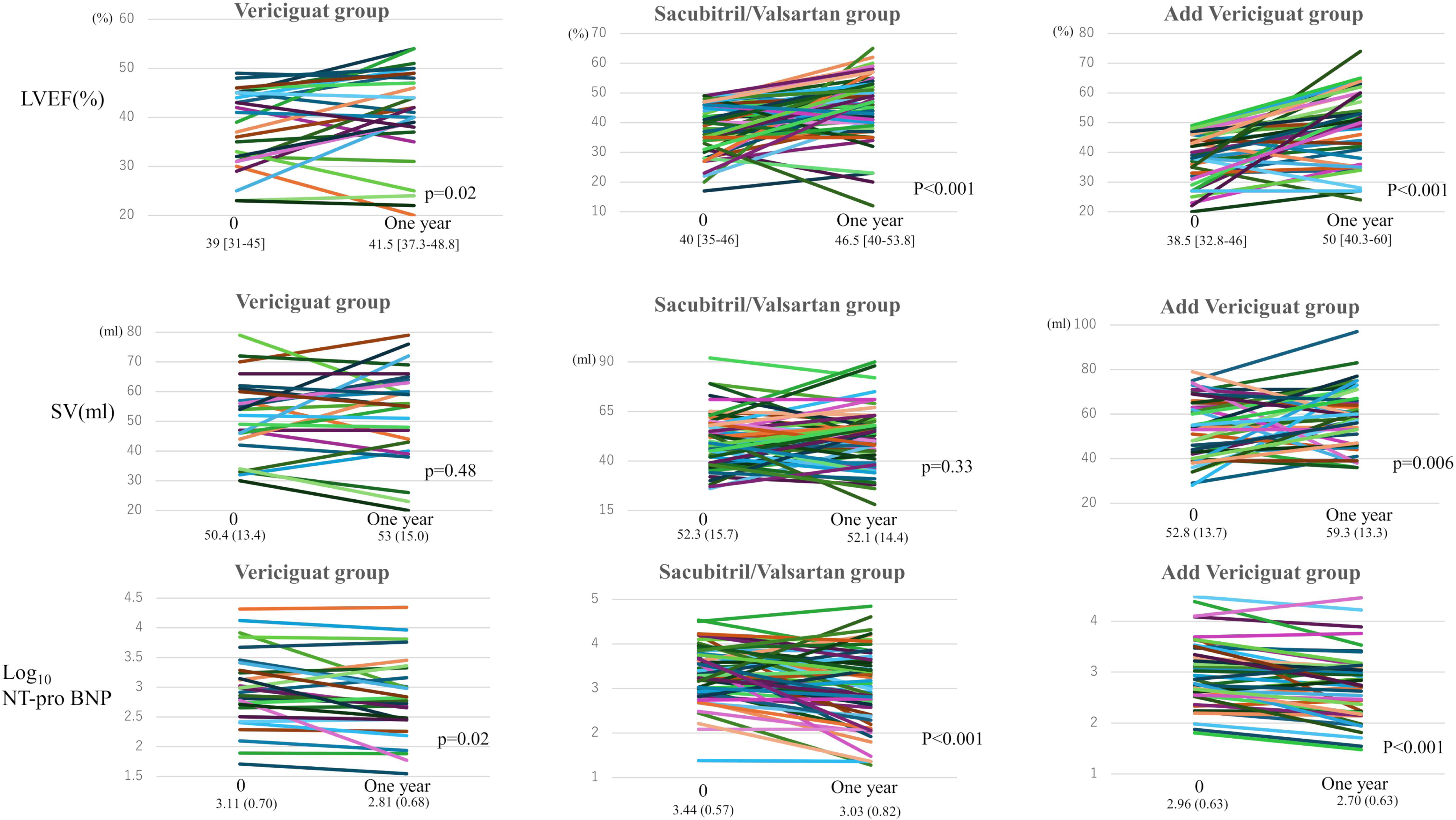
Changes in LVEF, SV, and Log_10_ NT-pro BNP 1 year post-initiation of target drug. In the vericiguat group, changes in LVEF, SV, and Log_10_ NT-pro BNP were measured 1 year post-initiation of vericiguat administration, while in the sacubitril/valsartan group, changes in LVEF, SV, and Log_10_ NT-pro BNP were measured 1 year after the initiation of sacubitril/valsartan administration. However, in the vericiguat group, sacubitril/valsartan had already been administered, and changes in LVEF, SV, and Log_10_ NT-pro BNP were measured 1 year after the initiation of vericiguat administration. Significant improvements were observed in LVEF and Log_10_ NT-pro BNP levels in all three groups. However, significant improvements in the SV were observed only in the vericiguat group.

**Figure 4:**
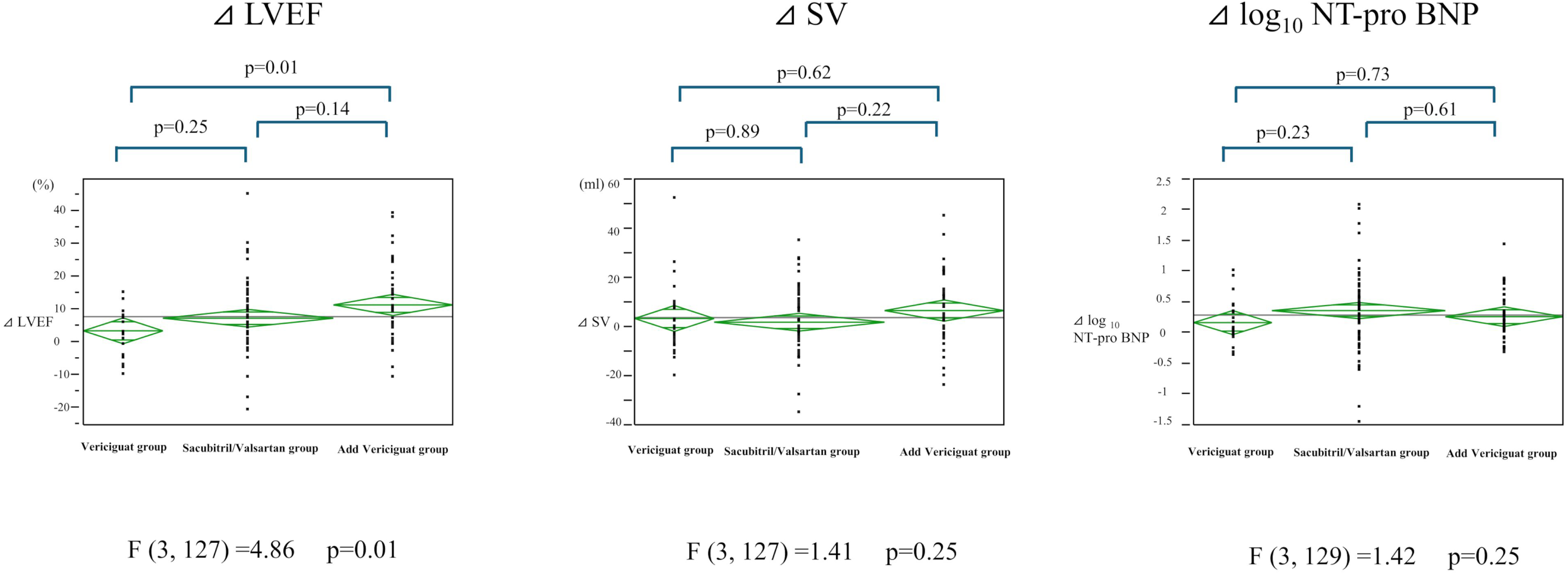
Three-group comparison of ΔLVEF, ΔSV, and ΔLog_10_ NT-pro BNP. The addition of vericiguat to sacubitril/valsartan did not result in significant improvements in LVEF, SV, or ΔLog_10_ NT-pro BNP. In this study, non-inferiority was also demonstrated between vericiguat monotherapy (with other heart failure medications other than sacubitril/valsartan) and sacubitril/valsartan monotherapy (with other heart failure medications other than vericiguat) in ΔLVEF, ΔSV, and ΔLog_10_ NT-pro BNP.

**Figure 5:**
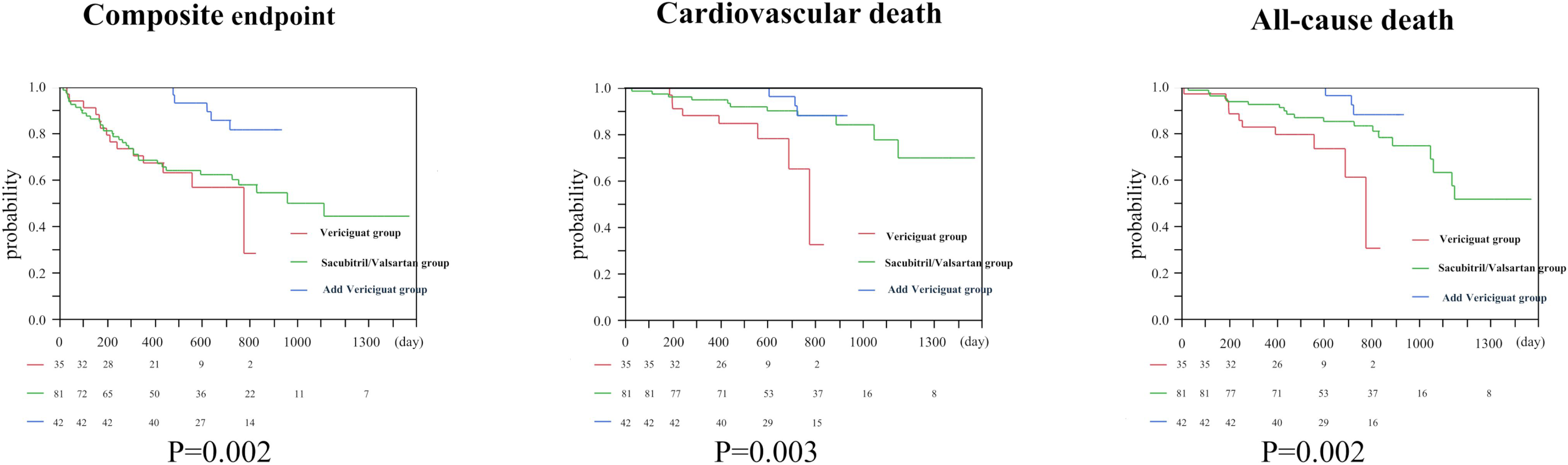
Kaplan–Meier curves and log-rank tests for composite, cardiovascular, and all-cause mortality. Log-rank tests showed significant differences among the three groups for the composite endpoint (p = 0.002), cardiovascular mortality (p = 0.003), and all-cause mortality (p = 0.002).

**Table 1.**
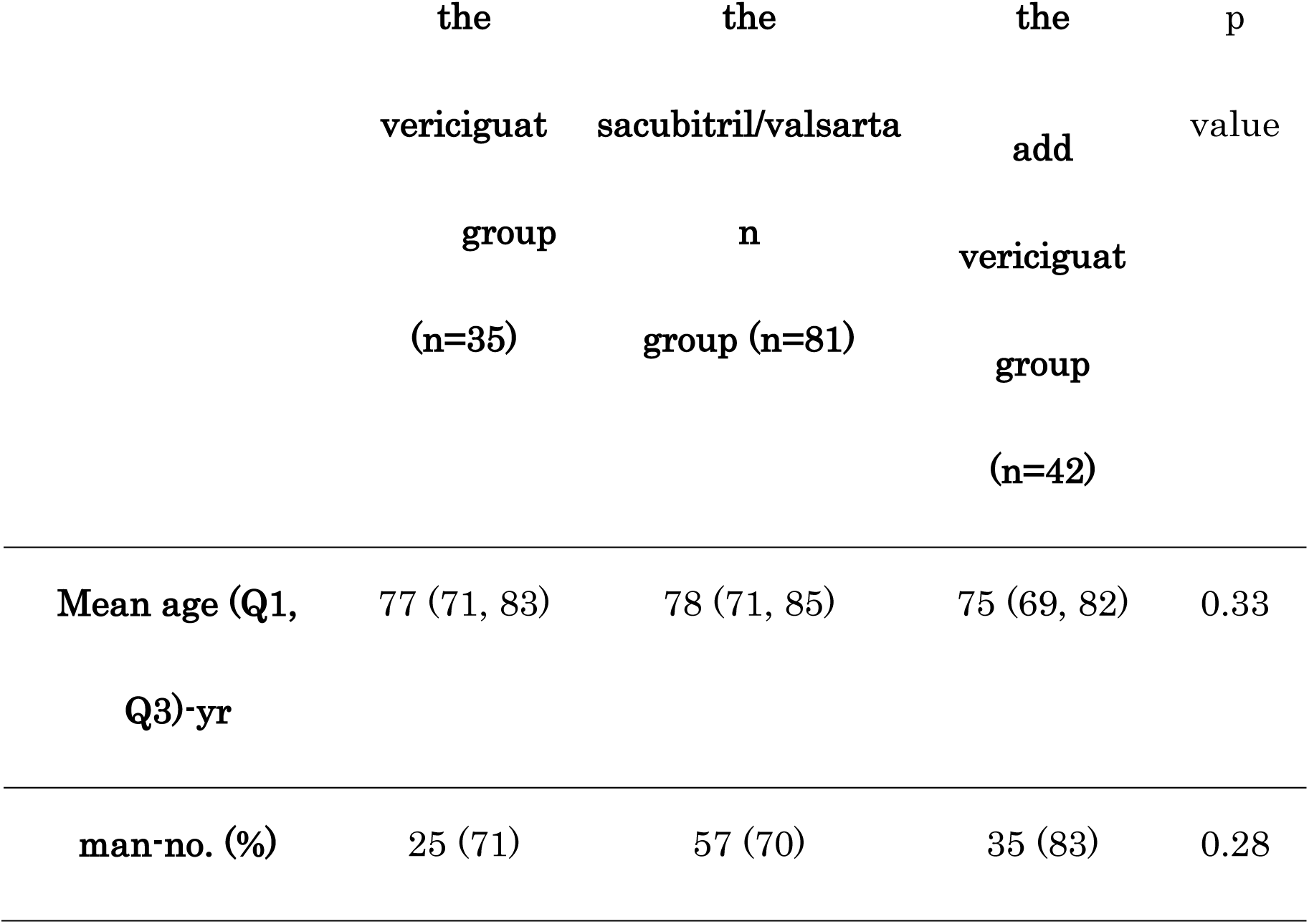
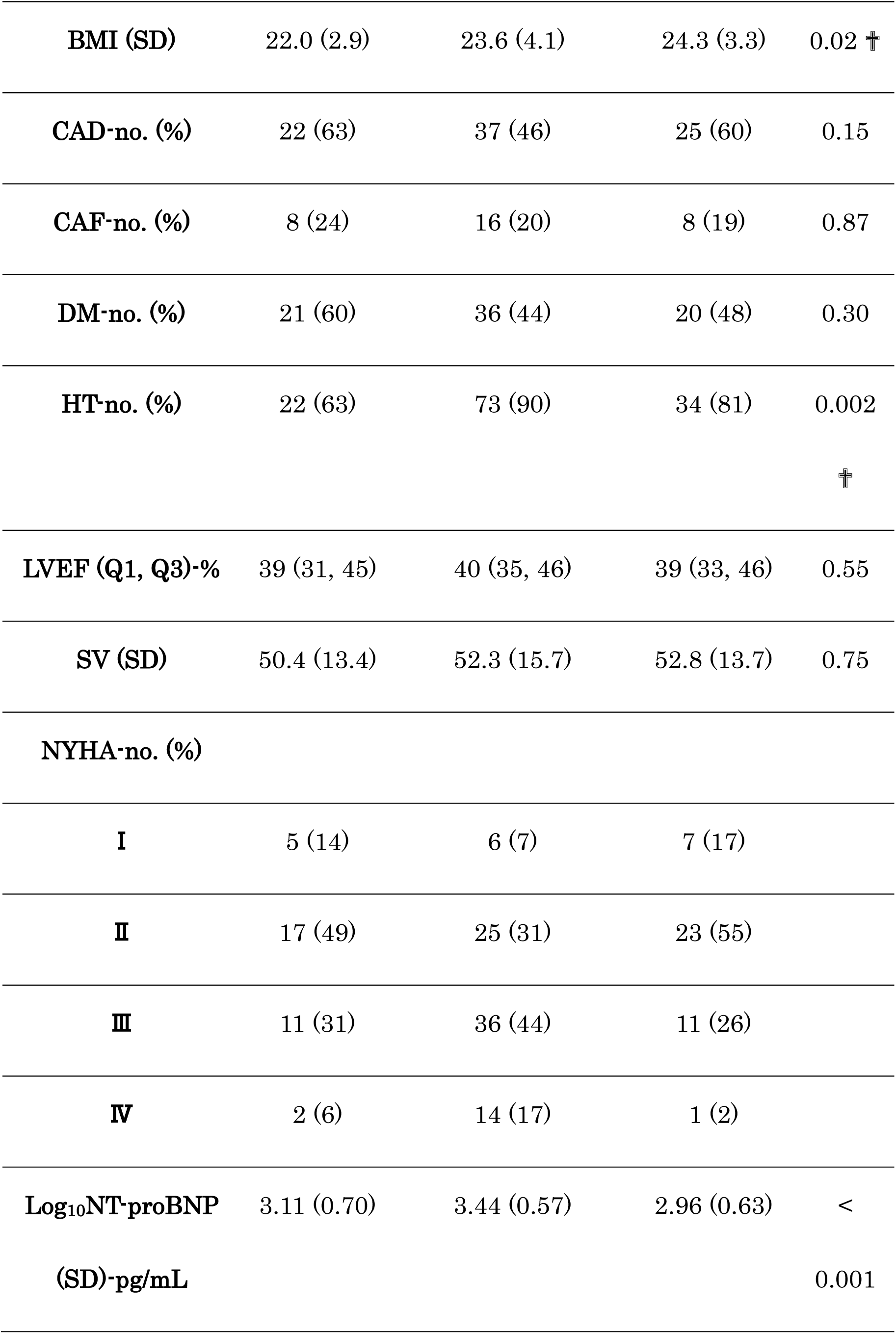
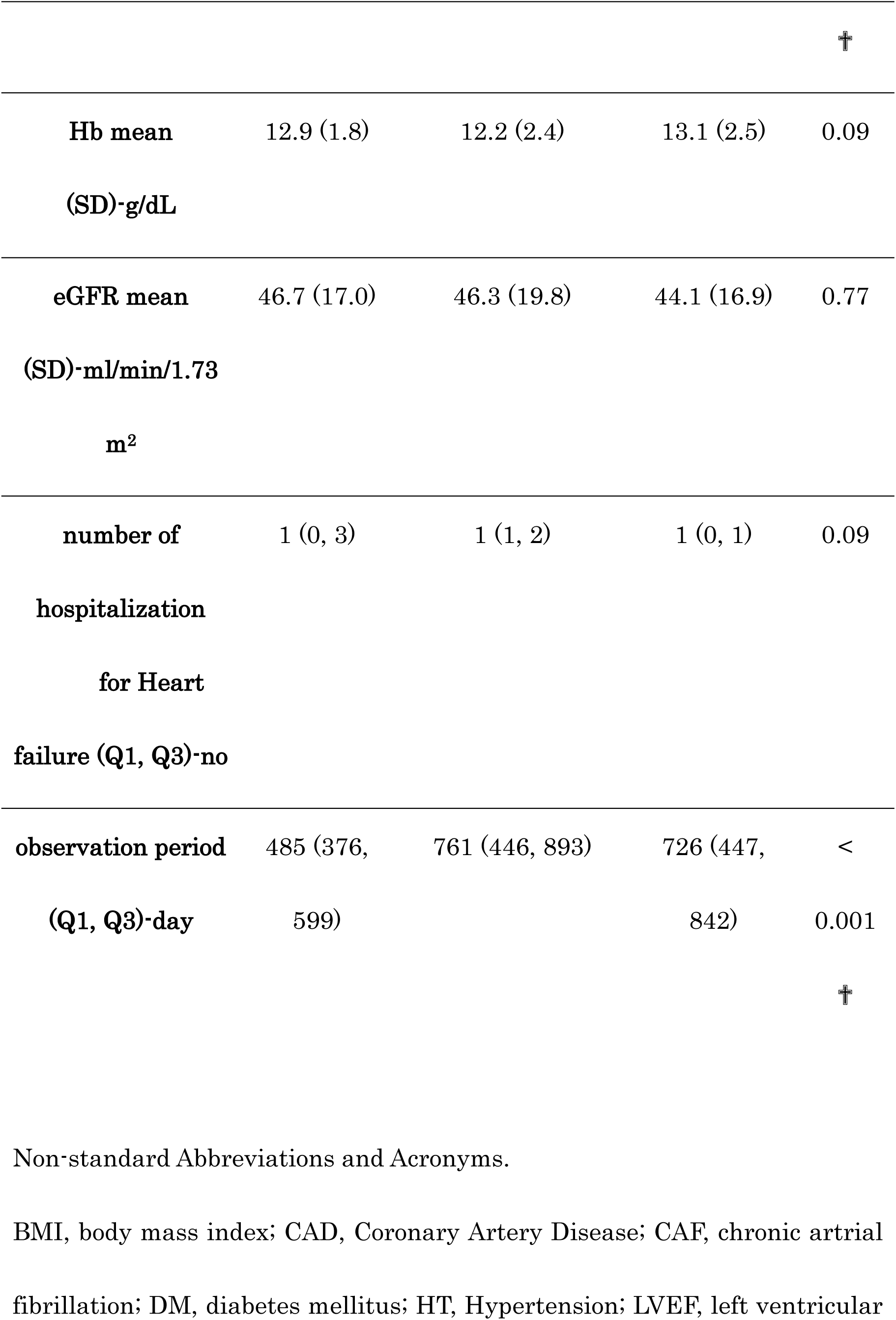
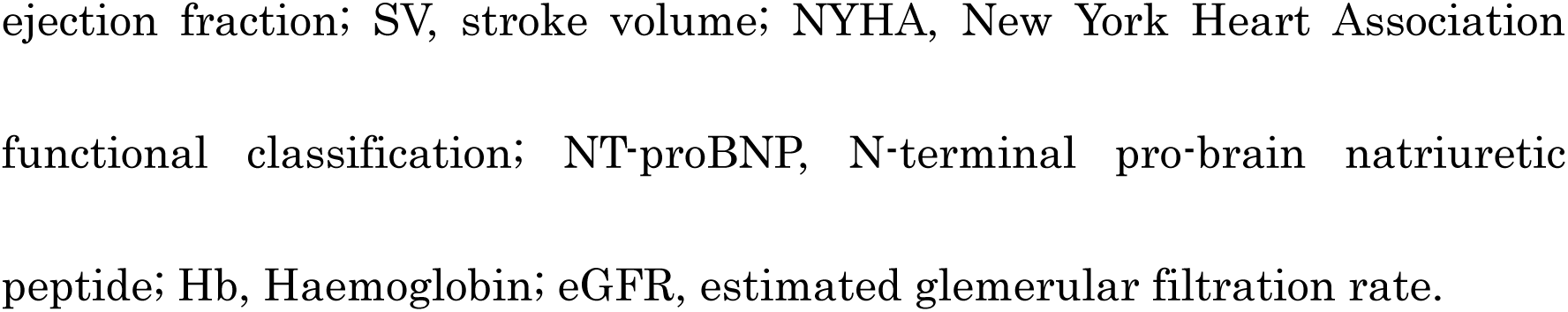
Baseline characteristics of the study population in the three groups (the vericiguat group, the sacubitril/valsartan group, and the add vericiguat group).

This retrospective, single-center cohort study analyzed patients treated with either sacubitril/valsartan or vericiguat between May 2021 and May 2024. Baseline echocardiographic evaluation was performed approximately 12 months (365 ± 90 days) after initiation of vericiguat or sacubitril/valsartan treatment in non-dialysis patients without HFpEF.

The study adhered to the principles of the Declaration of Helsinki and was approved by the Institutional Review Board of Kure Medical Center (approval No. 2023–43). Given the retrospective and opt-out design of the study, informed consent was obtained from all participants before data use. Eligible patients were divided into three treatment groups according to their therapeutic regimen:

(1) **vericiguat group**, comprising patients receiving vericiguat with standard heart failure therapies—including beta-blockers, MRAs, and sodium–glucose co-transporter 2 (SGLT2) inhibitors—but without sacubitril/valsartan; (2) **sacubitril/valsartan group**, consisting of patients receiving sacubitril/valsartan and other standard therapies without vericiguat; and (3) **add vericiguat group**, in which vericiguat was subsequently added to ongoing sacubitril/valsartan-based therapy.

Patients were excluded if they had discontinued sacubitril/valsartan or vericiguat at another institution, were on chronic dialysis, had cognitive impairment limiting adherence, or were unable to maintain treatment owing to insufficient family support. Patients with preserved LVEF ≥50% were also excluded from the study. Furthermore, when evaluating longitudinal changes in echocardiographic and biomarker parameters, patients were excluded if the baseline echocardiogram was obtained >3 months before the initiation of sacubitril/valsartan or vericiguat or if the follow-up echocardiogram was performed within 150 days or beyond 2 years after drug initiation. This restriction ensured that both baseline and 1-year follow-up data accurately reflected the treatment period, avoiding cases in which the timing of echocardiography was too distant to represent the pre-treatment or 1-year post-treatment status.

The initiation and titration of each medication followed the guideline-based dosing strategies determined by the treating cardiologist. Sacubitril/valsartan was initiated during either outpatient care or hospitalization and uptitrated to a target daily dose of 100–400 mg. Vericiguat was started at 2.5 mg daily and increased to a maximum of 10 mg every 1–3 months, depending on blood pressure tolerance. Dose adjustments or delays were made in cases of symptomatic hypotension, such as dizziness or lightheadedness.

Baseline characteristics, including demographic, laboratory, echocardiographic, and medication data, were obtained from electronic health records. Echocardiographic examinations were performed by a board-certified sonographer (Japan Society of Echocardiography) who was blinded to the patient treatment allocation. LVEF and SV were measured using two-dimensional echocardiography.

Statistical analyses were conducted using JMP software (version 9.0; SAS Institute, Cary, NC, USA). The distribution of continuous variables was assessed for normality. Depending on variance equality, comparisons between groups were performed using one-way analysis of variance (ANOVA), Student’s t-test, or Wilcoxon rank-sum test. Categorical variables were analyzed using the chi-square or Fisher’s exact tests. Tukey’s post-hoc test was used for multiple comparisons among the three groups.

Time-to-event outcomes for cardiovascular and all-cause mortality, and composite cardiovascular events were evaluated using Cox proportional hazards regression models, yielding hazard ratios (HRs) with 95% confidence intervals (CIs). Statistical significance was defined as a two-sided p-value <0.05.

Efficacy analyses were performed for patients who were able to continue receiving sacubitril/valsartan and vericiguat, and were followed up.

## Results

Thirty-five patients received vericiguat in combination with other heart failure medications, excluding sacubitril/valsartan (vericiguat group; four of 39 initially enrolled patients withdrew). Another 127 patients were treated with sacubitril/valsartan plus standard therapies such as beta-blockers, MRAs, and SGLT2 inhibitors (sacubitril/valsartan group; seven of 134 patients withdrew). Among them, 42 patients later received vericiguat added to ongoing sacubitril/valsartan therapy (add vericiguat group; 4 of the 46 initially enrolled patients withdrew) (Figure 1).

The median age was 77 years (IQR: 71–83 years) in the vericiguat group, 78 years (IQR: 71–85 years) in the sacubitril/valsartan group, and 75 years (IQR: 69–82 years) in the vericiguat group, with no significant intergroup differences (p = 0.33). Dose adjustment, downtitration, or discontinuation of sacubitril/valsartan or vericiguat was performed at the discretion of the treating cardiologist based on the patient’s blood pressure and reported symptoms, such as dizziness or fatigue. The median follow-up periods from the initiation of sacubitril/valsartan or vericiguat therapy were 455 days (IQR: 376–599 days) in the vericiguat group, 761 days (IQR: 446–893 days) in the sacubitril/valsartan group, and 726 days (IQR: 447–842 days) in the vericiguat group. Baseline characteristics differed significantly among the three groups in terms of body mass index (BMI), prevalence of hypertension, and log-transformed NT-proBNP values (BMI, p = 0.02; hypertension, p = 0.002; log_10_ NT-proBNP, p < 0.001) (Table 1). The use of diuretics and heart failure medications other than vericiguat and sacubitril/valsartan was introduced in each group, and the simple Guideline-Directed Medical Therapy (GDMT) scores^8^ are shown in Figure 2. Furthermore, for each of the three groups, LVEF, SV, and Log_10_ N-terminal pro-B-type natriuretic peptide (NT-pro BNP) at induction (the target drug for the vericiguat group was vericiguat, for the sacubitril/valsartan group was sacubitril/valsartan, and for the add vericiguat group was vericiguat) and change at 1 year (365 ± 90 days) by Wilcoxon’s rank sum test (LVEF and SV) or t-test (Log_10_ NT-pro BNP) (Figure 3). The LVEF and Log_10_ NT-pro BNP levels significantly improved in all groups. Next, a t-test was performed for changes in the LVEF (ΔLVEF%), SV (ΔSV mL), and Log_10_ NT-pro BNP (ΔLog_10_ NT-pro BNP pg/mL) between the three groups at induction and 1 year later, and found significant differences in ΔLVEF (p = 0.01). The Honest Significant Difference (HSD) test revealed significant differences between the vericiguat and vericiguat groups, but not between the vericiguat and sacubitril/valsartan groups (Figure 4).

This study analyzed the prognosis of three items: ① composite endpoint (combination of cardiovascular mortality, emergency hospitalization due to heart failure, and emergency consultation due to heart failure), ② cardiovascular mortality, and ③ all-cause mortality.

Cumulative event rates were estimated using the Kaplan–Meier method for the three groups. The event rates at 12 months were as follows: ① for the composite endpoint, 32.6%, 31.4%, and 0% in the vericiguat, sacubitril/valsartan, and added vericiguat groups, respectively; ② for cardiovascular mortality, 12%, 5%, and 0% in the vericiguat, sacubitril/valsartan, and added vericiguat groups, respectively; and ③ for all-cause mortality, 18%, 7%, and 0% in the vericiguat, sacubitril/valsartan, and vericiguat groups, respectively. The log-rank test revealed significant differences among the three groups for composite endpoint, cardiovascular mortality, and all-cause mortality (composite endpoint, p = 0.002; cardiovascular mortality, p = 0.003; and all-cause mortality, p = 0.002) (Figure 5).

As reported in the EJHF, multivariate analysis was performed using a Cox proportional hazards model with HT, BMI, Log_10_ NT-pro BNP, and treatment group (vericiguat, sacubitril/valsartan, and vericiguat) as covariates. The results showed that the vericiguat group showed a significant difference in the composite endpoint (composite of cardiovascular mortality, emergency hospitalization due to heart failure, and emergency consultation due to heart failure) compared with the sacubitril/valsartan group.^7^

## Discussion

This study is unique because it evaluated the additional effects of vericiguat after initiating sacubitril/valsartan treatment in all patients. In contrast, the initiation rates of sacubitril/valsartan in previously reported large-scale randomized controlled trials (RCTs), the VICTORIA and VICTOR trials, were only 14.5% and 56%, respectively.^4,9^ Therefore, our cohort represents a real-world population that accurately reflects contemporary GDMT practices.

Furthermore, the median NT-proBNP level in our cohort (1824 pg/mL; log_10_ ≈ 3.26) was intermediate between that reported in VICTORIA (2816 pg/mL) and VICTOR (1375 pg/mL). Furthermore, the study included patients with NT-pro BNP levels of 6000 pg/mL or higher, including patients who were currently hospitalized for heart failure or had a recent history of hospitalization for heart failure. This indicates that this study’s participants were a typical population in current clinical settings with moderately severe heart failure. Furthermore, by incorporating log_10_ NT-proBNP as a covariate in the multivariate Cox proportional hazards analysis, the effect of disease severity was statistically corrected before evaluating prognosis, and the observed results were not simply due to differences in baseline disease severity. Based on these findings, this study is considered significant as it provides complementary data that bridges the evidence provided by RCTs to clinical practice.

Vericiguat is a novel drug for chronic heart failure that targets the nitric oxide (NO)-soluble guanylate cyclase (sGC)-cyclic guanosine monophosphate (cGMP) pathway (NO pathway), consequently increasing cGMP production by increasing the sensitivity of sGC to NO or by directly stimulating sGC in an NO-independent manner.

Besides the NO pathway, the natriuretic peptide-mediated pathway (ANP pathway) is involved in intracellular cGMP production. These pathways act on sGC and particulate guanylate cyclase(pGC) to activate cGMP-dependent protein kinase G (PKG). These pathways also act on vascular smooth muscle cells and contribute to the phosphorylation of titin and myosin-binding protein C, which are involved in myocardial contraction and relaxation, thereby improving myocardial function.^10,11^ The NO and ANP pathways are believed to exist in functionally and structurally distinct compartments. The cGMP produced by the NO pathway is located in a compartment controlled by phosphodiesterase-5 (PDE-5), whereas that produced by the ANP pathway is located in a compartment controlled by phosphodiesterase-9 (PDE-9). PDE-5 and PDE-9 are considered to have different intracellular localizations and effects on cGMP levels.^12^

Additionally, in a mouse heart failure model, it has been reported that therapeutic drugs targeting the ANP (PDE-5 inhibitor) and NO (PDE-9 inhibitor) pathways similarly activate the cGMP pathway and contribute to the suppression of remodeling. However, these drugs affect different downstream genes, with only 10% of the genes interacting in common, indicating different behaviors depending on the activity of each pathway.^13^ In this real-world study, the addition of vericiguat to sacubitril/valsartan did not further improve LVEF compared to sacubitril/valsartan alone. However, patients receiving the combination therapy showed greater improvement in LVEF than those treated with vericiguat alone (Figure 4). When comparing the three groups (vericiguat, sacubitril/valsartan, and add-vericiguat), the change in LVEF (ΔLVEF) did not differ significantly between the vericiguat and sacubitril/valsartan groups (p = 0.25). Nevertheless, both groups showed significant improvements from baseline to 1 year (vericiguat group, p = 0.02; sacubitril/valsartan group, p < 0.001) (Figure 3). These findings suggest that vericiguat provides LVEF benefits comparable with those of sacubitril/valsartan, supporting its potential as an alternative therapeutic option for patients who cannot tolerate or maintain angiotensin receptor-neprilysin inhibitor (ARNI) therapy.

Notably, as shown in Figure 5, the Kaplan–Meier curves for the vericiguat and sacubitril/valsartan groups for the composite endpoint followed similar curves until approximately day 600, whereas the courses of cardiovascular and all-cause mortality in the Kaplan–Meier curves diverged from approximately day 200. If the composite endpoint is considered to represent the occurrence of events during the transition from stage B to stage C heart failure, cardiovascular mortality, and all-cause mortality can be considered to represent the occurrence of events during the transition from the end of stage C heart failure to stage D heart failure. The Kaplan–Meier curves for the vericiguat and sacubitril/valsartan groups for the composite endpoint were consistent, suggesting that vericiguat can be as effective as sacubitril/valsartan in preventing progression from stage B to stage C. However, the Kaplan–Meier curves for cardiovascular and all-cause mortality showed a divergence between the vericiguat and sacubitril/valsartan groups from around day 200, suggesting that there may be a limit to the ability to sustain the condition once it reaches end-stage C or D. On the other hand, sacubitril/valsartan may have the ability to slow the progression of heart failure even when it reaches end-stage C or D.

This study has several limitations. First, this was a single-center retrospective observational analysis with a relatively small sample size (n = 158), which may have limited the generalizability of the findings. Second, the duration of follow-up differed among patients owing to variability in the timing of drug initiation. Third, the analysis included only patients who were able to continue both sacubitril/valsartan and vericiguat, while those who discontinued therapy, transferred care to other institutions, or had difficulty maintaining treatment (e.g., due to cognitive decline) were excluded. Therefore, selection bias cannot be fully excluded, and the results should be interpreted with caution.

Future prospective studies with larger sample sizes and longer follow-up periods are warranted to validate these findings and clarify the long-term effects of continuous dual therapy.

Nevertheless, this real-world analysis provides valuable insights into the clinical course of patients treated with contemporary heart failure pharmacotherapy outside the strict setting of RCTs.

For HFrEF, quadruple therapy and other comprehensive combination regimens, including ARNIs, are generally recommended.^14^ When ARNI is not feasible, clinicians often rely on alternative RAAS-based combinations. In this real-world study of non-dialysis patients without HFpEF, vericiguat demonstrated potential as a substitute therapeutic option for ARNIs in terms of LVEF improvement. This study also demonstrated that adding vericiguat to sacubitril/valsartan before the onset of refractory or fatal heart failure may delay the progression to heart failure requiring hospitalization. These findings suggest that strategic sequencing and early incorporation of vericiguat could optimize the continuum of medical therapy in heart failure with nonpreserved ejection fraction patients.

## Abbreviations (Alphabetical order)

ANOVA: Analysis of variance
ARNI: Angiotensin receptor–neprilysin inhibitor
BMI: Body mass index
CI: Confidence interval
cGMP: Cyclic guanosine monophosphate
EJHF: European Journal of Heart Failure
GDMT: Guideline-directed medical therapy
HFpEF: Heart failure with preserved ejection fraction
HFrEF: Heart failure with reduced ejection fraction
HR: Hazard ratio
HSD: Honest Significant Difference
LVEF: Left ventricular ejection fraction
MRA: Mineralocorticoid receptor antagonist
NT-proBNP: N-terminal pro-B-type natriuretic peptide
PDE-9: Phosphodiesterase-9
pGC: Particulate guanylate cyclase
PKG: Protein kinase G
RCTs: Randomized controlled trials
sGC: Soluble guanylate cyclase
SGLT2: Sodium–glucose co-transporter 2
SV: Stroke volume

## Acknowledgements

The authors acknowledge Akita Tomoyuki, MD, PhD, Department of Epidemiology and Infectious Disease Control, Graduate School of Biomedical Sciences, Hiroshima University for analysis. The authors acknowledge Ms. Yoshimi Shitakubo, Department of Clinical Research Department, NHO Kure Medical Center for analysis. The authors acknowledge Ms. Aya Ohshita, medical administrative assistant, NHO Kure Medical Center for data acquisition. The authors would like to thank Editage (www.editage.jp) for English language editing.

## Data Availability

The data that support the findings of this study are available from the corresponding author upon reasonable request.

